# Automated Melanoma Screening: A Machine Learning Pipeline for Mole Detection, Boundary Segmentation, and ABCD(E) Feature Extraction

**DOI:** 10.64898/2026.06.29.26356601

**Authors:** Mahla Abdolahnejad, Emily Pascazi, Madison Lee, Johnson Cheng, Faith Poon, Manuella Kyeremeh, Hannah O. Chan, Rakesh Joshi, Colin Hong

## Abstract

Early detection of suspicious moles remains the most effective means of reducing mortality from skin cancer, yet systematic screening is constrained by the time and expertise required for manual mole assessment. This paper presents an end-to-end computational pipeline that utilizes wide-angle skin photographs (including consumer-grade smartphone images) and produces quantitative ABCD (Asymmetry, Border irregularity, Color variegation, Diameter) feature scores for every detected mole. The pipeline operates in four stages: mole detection via adaptive thresholding and blob analysis, super-resolution enhancement using EDSR, false-positive filtering using a brightness-based statistical criterion, and lesion segmentation using the Boundary Attention Mapper (BAM). BAM generates high-resolution segmentation masks by fusing early-layer activations with GradCAM heatmaps from a trained EfficientNet-B7 classifier, achieving 90.45% accuracy on the ISIC2017 dataset, outperforming both conventional GradCAM (87.78%) and dedicated segmentation architectures, including DeepLabv3 and SAM v2 by more than 5 percentage points in Dice score. The EfficientNet-B7 backbone achieves a micro-average AUC of 0.97 across eight lesion classes, with a melanoma AUC of 0.99. Color quantification uses K-means clustering with a threshold calibrated on the PH2 dataset (MSE = 1.425). Applied to 87 wide-angle images, the mole detection module achieved an F1 score of 86%. The system outputs a structured CSV of per-lesion ABCD scores suitable for clinical triage and longitudinal tracking. A clinical validation study with dermatologists and surgeons is underway to assess concordance between automated and expert assessments.

## 1. Introduction

Melanoma has the highest mortality rate among the most common skin cancers. It accounts for a disproportionate share of skin cancer deaths despite comprising a minority of diagnoses [1], and its incidence has climbed steadily across fair-skinned populations with cumulative ultraviolet exposure [2]. The clinical stakes are usually stage-dependent: five-year survival exceeds 98% when the disease is early and confined to the skin, but drops below 25% once it has metastasized [3]. The gap between a curable local lesion and a largely fatal systemic disease is the entire rationale for early detection. Early detection of suspicious moles spares patients from aggressive surgical and systemic therapies, reduces downstream healthcare costs, and aligns with the broader shift toward preventive medicine [4].

The standard clinical tool for detecting lesions early is total-body mole mapping: a systematic photographic survey of the skin surface that creates a baseline record of every pigmented lesion [5]. For high-risk patients, namely those with a family history of melanoma, large numbers of atypical nevi, or immunosuppressive conditions [6, 7], mapping is most valuable as it allows clinicians to spot new or changing moles across successive visits. When combined with dermoscopy, mole mapping can reveal malignant changes that are invisible to the unaided eye [8]. But the process is slow. Manual lesion documentation averages 14.1 minutes per patient during a baseline exam [9], and this time cost limits the number of patients a clinic can screen in a day. Automation is not merely convenient here; it is the difference between a screening tool that works at scale and one that does not.

When a clinician evaluates an individual mole, the assessment typically follows the ABCDE rule: Asymmetry, Border irregularity, Color variation, Diameter greater than 6 mm, and Evolution [10]. These criteria are used to teach trainees and structured enough to translate into computable features, which is why the ABCD rule has become a backbone of computer-aided diagnosis (CAD) systems as well as bedside dermatology [11]. Asymmetry captures uneven shape or pigment distribution; border irregularity flags jagged or poorly defined edges; color variegation denotes mixed shades of brown, black, red, or white; and diameter provides a size-based risk threshold. Clinicians add a fifth criterion, Evolving, to flag dynamic changes over time [7]. The rule is a heuristic, not a definitive diagnostic, but it provides a quantifiable starting point for distinguishing moles that require biopsy from those that do not.

Automating the ABCD rule for wide-angle skin photographs is more challenging than it appears. Most deep learning systems for lesion analysis have been trained on high-quality dermoscopic images taken under controlled lighting with standardized equipment [13]. Smartphone photographs vary in resolution, white balance, angle, and background clutter [12], and those variations degrade model performance, especially across different skin tones [14]. Segmentation is the critical bottleneck: if the algorithm cannot accurately delineate the boundary between the mole and the skin, any downstream ABCD measurement inherits that error. Asymmetry, border irregularity, and diameter are all computed directly from the segmentation boundary, so even small inaccuracies in boundary detection propagate into unreliable feature scores [15].

This paper presents an end-to-end pipeline for automated mole detection, segmentation, ABCD feature extraction, and suspicion scoring from wide-angle skin images. The system is designed to work with consumer-grade smartphone photographs, not just clinical dermoscopy, making it applicable to teledermatology and home-monitoring scenarios. At its core, the pipeline uses the Boundary Attention Mapper (BAM), a saliency-based segmentation method that combines early-layer activation maps from a trained CNN with GradCAM heatmaps to produce high-resolution lesion masks. For color quantification, we pair K-means clustering with a threshold calibrated using expert-annotated ground truth data. The result is a system that takes a smartphone photo as input and returns a structured table of ABCD scores for every mole in the image.

## 2. Methodology

### 2.1. System Overview

The pipeline has four stages, illustrated in Figure 1. A wide-angle skin photograph is entered; per-lesion ABCD feature scores are derived. Stage one (**Mole Detection**) localizes individual lesions using adaptive thresholding and blob detection. Stage two (**Image Processing**) enhances each cropped mole patch through 4× super-resolution upsampling. Stage three (**False Positive Reduction**) discards spurious detections via a brightness-based filter. Stage four (**Segmentation and ABCD Extraction**) segments each validated mole using BAM and computes the four ABCD features from the resulting mask.

**Figure 1:**
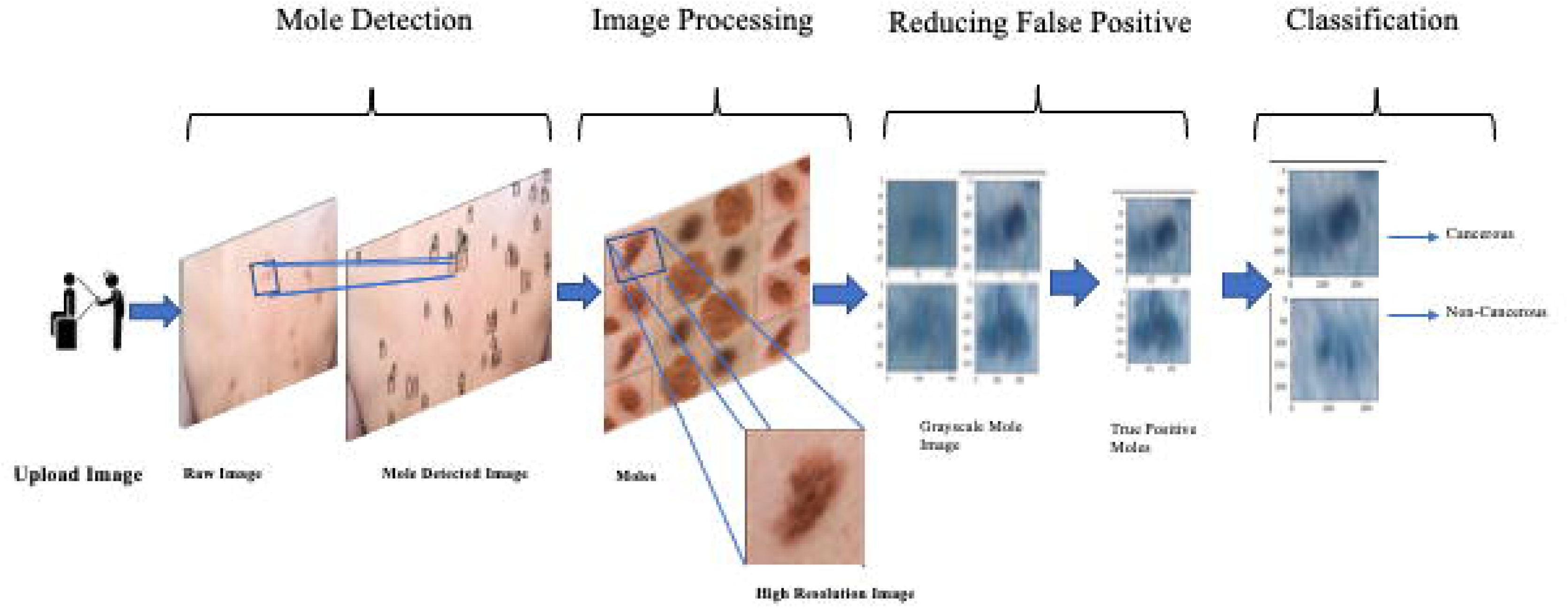
Overall system pipeline for automated mole detection and ABCD analysis from wide-angle skin imagery.

### 2.2. Mole Detection

The detection module takes a full wide-angle image and returns bounding boxes around individual moles (Figure 2). It proceeds in three steps: pre-processing, adaptive thresholding, and blob detection.

**Figure 2.**
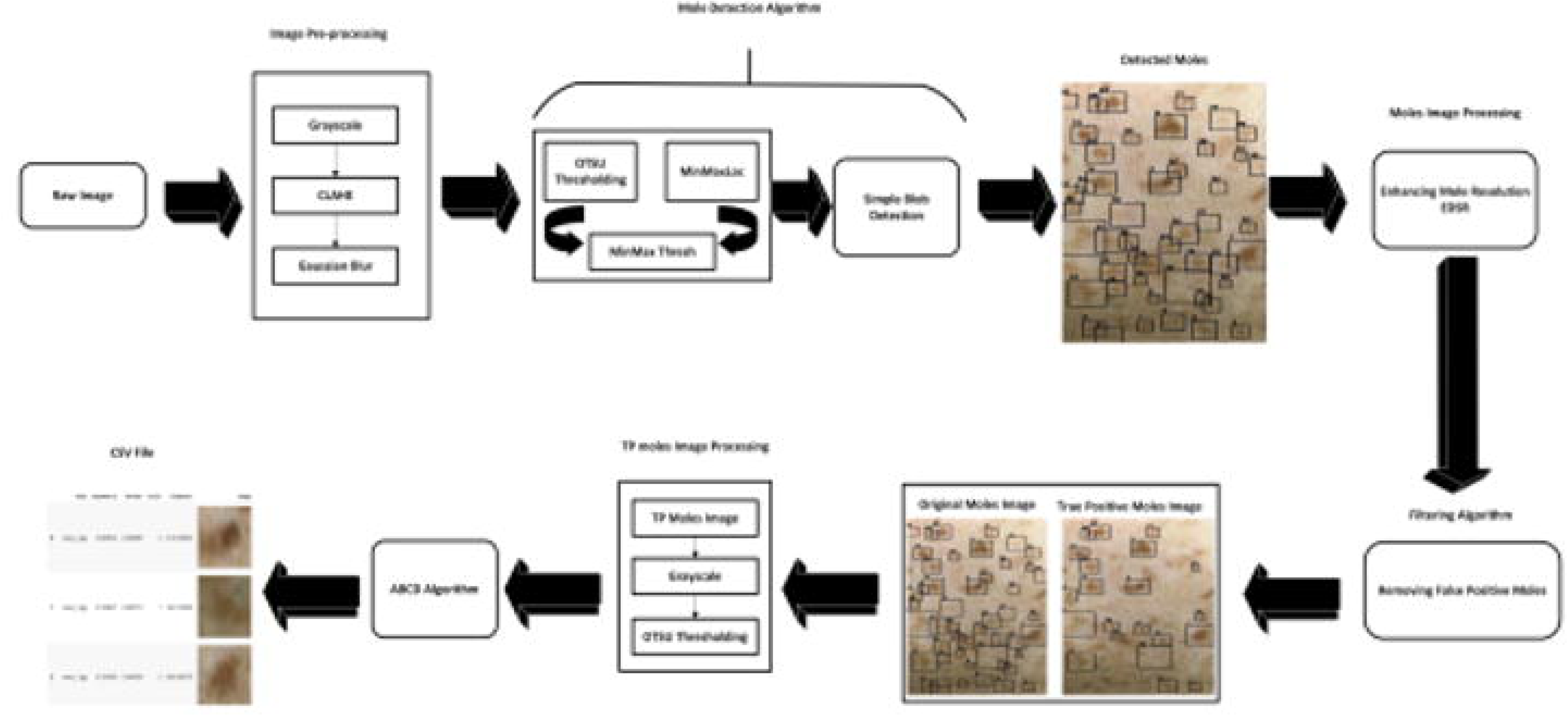
Mole Mapping System Architecture.

#### 2.2.1. Image Pre-processing

Each image is converted to greyscale, then contrast-normalized using Contrast Limited Adaptive Histogram Equalization (CLAHE) with a clip limit of 0.01 and tile grid of 15 × 15. The CLAHE step compensates for the uneven illumination, skin curvature, and shadowing typical of wide-angle photography. A 15 × 15 Gaussian blur (σ computed from kernel size) is applied next to suppress skin texture and fine-grained noise while preserving the spatial extent of mole regions.

#### 2.2.2. Adaptive Thresholding

Threshold bounds for the blob detector are set through a dual-method strategy. The blurred greyscale image is inverted and processed with Otsu’s method to obtain *T*_OTSU. Separately, *minMaxLoc* extracts the global minimum (*T*_min) and maximum (*T*_max) intensities from the non-inverted image. The final bounds are:

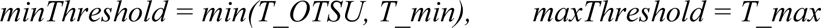

Neither Otsu’s method nor raw intensity extrema alone handles the full range of wide-angle lighting conditions. Their combination provides coverage across bright, dark, and mixed-exposure images.

#### 2.2.3. Blob Detection and Mole Extraction

Moles are detected using OpenCV’s SimpleBlobDetector with *blobColor* = 0, area filtering between 10 and 2,000 pixels, and all shape filters (circularity, convexity, inertia) disabled. The area bounds exclude sub-pixel noise at the low end and large non-mole structures (birthmarks, tattoos) at the high end. Each detected keypoint yields a square crop of side *w* = 2 × *keypoint.size*, taken from the original color image (Figure 3).

**Figure 3.**
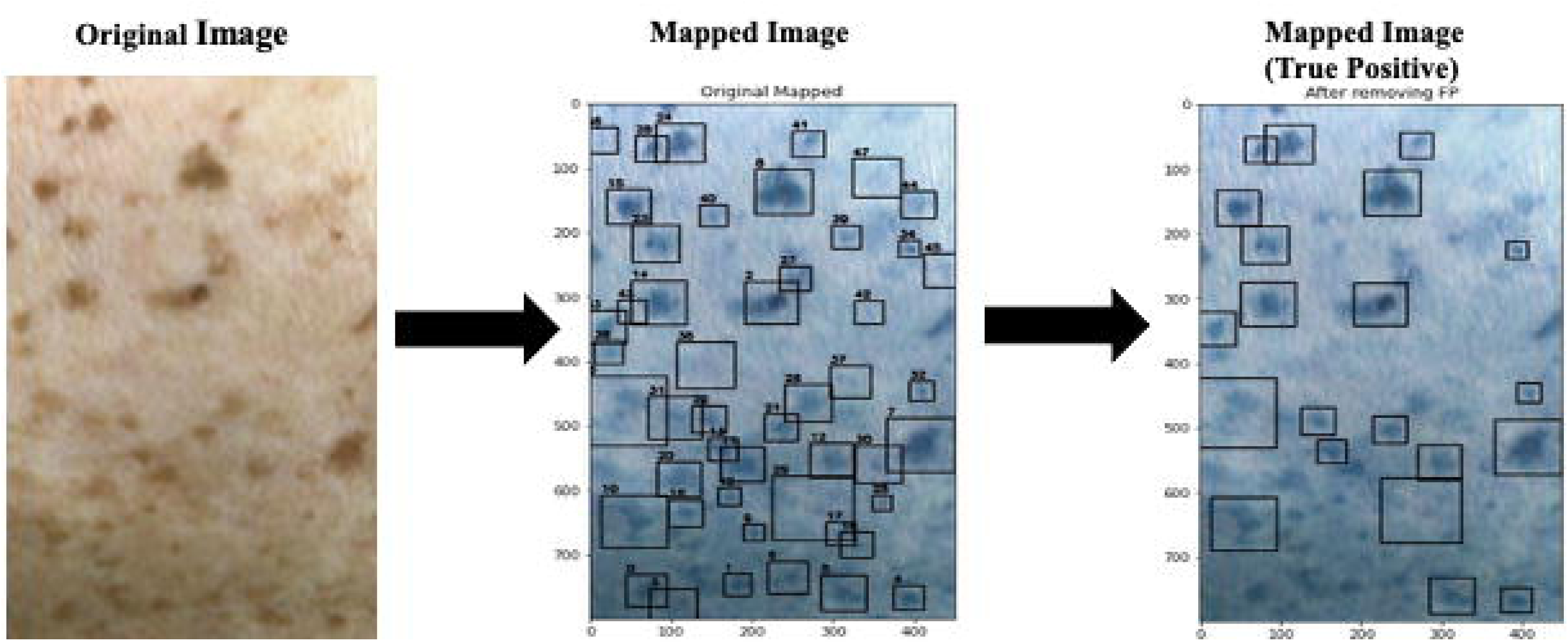
Mole detection and False positive Removal.

#### 2.2.4. Super-Resolution Enhancement

Moles in wide-angle images often span only a handful of pixels. Each crop is upsampled 4× using the pre-trained Enhanced Deep Super-Resolution (EDSR) model, accessed through OpenCV’s cv2.dnn_superres module [16]. The upsampling recovers texture and boundary detail that would otherwise be lost in the downstream segmentation and feature extraction stages.

#### 2.2.5. False Positive Removal

Not every detected blob is a mole. Skin folds, hair follicles, and imaging artifacts produce false positives that need filtering. The key observation is that genuine moles have a dark core against lighter surrounding skin, whereas false positives tend to be uniformly bright.

For each enhanced patch, the greyscale image is analyzed to extract the three most frequent pixel intensities (*m*₁, *m*₂, *m*₃) via successive mode computation. An amplification factor *r* is assigned: 1.2 if all three modes exceed 200 or fall below 100 (bright or dark images, respectively), and 1.3 otherwise. A detection is retained as a true positive only if *r* × min(*I*) < mean(*I*), confirming that the darkest region is sufficiently darker than the patch mean, the hallmark of a pigmented lesion against a skin background (Figure 3, right panel).

### 2.3. Lesion Segmentation

Accurate boundary delineation is the single most important prerequisite for reliable ABCD scoring. We use the Boundary Attention Mapper (BAM), which repurposes the internal representations of a trained CNN classifier to generate segmentation masks without requiring a dedicated segmentation network.

#### 2.3.1. EfficientNet-B7 Lesion Classifier

The backbone is a pre-trained EfficientNet-B7 [17] accepting 600 × 600 × 3 inputs. Its architecture begins with a convolutional layer producing 64 maps at 300 × 300, followed by seven MBConv blocks of progressively decreasing resolution and increasing depth: Block 1 (300×300×32), Block 2 (150×150×48), Block 3 (75×75×80), Block 4 (38×38×160), Block 5 (38×38×242), Block 6 (19×19×384), Block 7 (19×19×640), and a final convolutional block (19×19×2560). Global average pooling reduces this to a 1×2560 vector for multi-class diagnosis across eight categories: basal cell carcinoma (bcc), benign keratosis-like lesions (bkl), dermatofibroma (dn), melanoma (mel), melanocytic nevi (nv), other (other), squamous cell carcinoma (scc), and vascular lesions (vasc). The architecture is shown in Figure 4.

**Figure 4.**
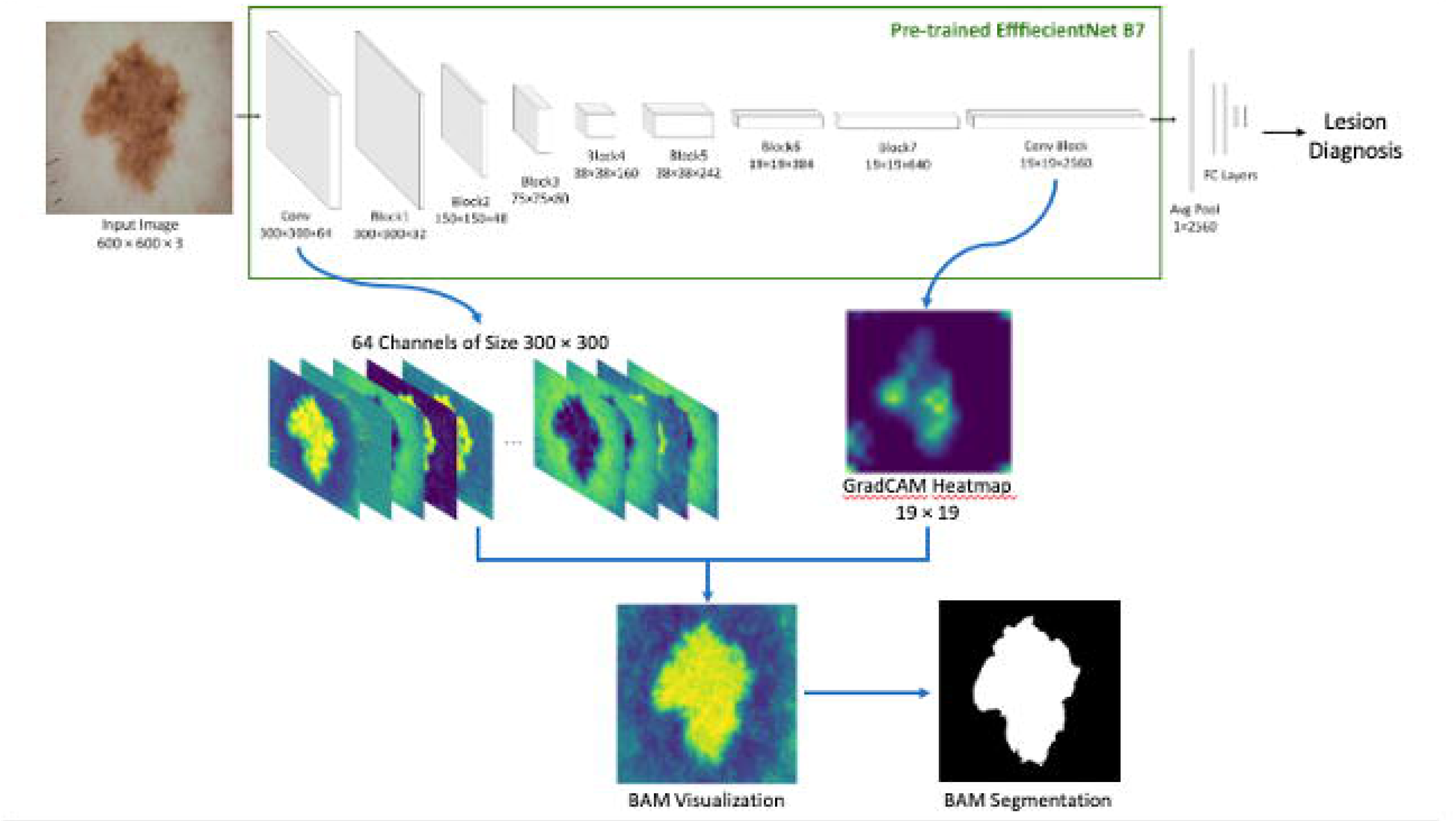
Architecture of the Boundary Attention Mapper (BAM) method (modified from Abdolahnejad et al., 2025)

The classifier was trained on a dataset of 36,000 smartphone-captured skin lesion images, comprising both original and augmented samples. Data augmentation included standard geometric and photometric transformations to improve generalization across the imaging variability inherent in consumer-grade photographs. The dataset was partitioned into training (70%), validation (20%), and test (10%) splits. Training proceeded with cross-entropy loss and standard hyperparameter tuning on the validation set; the results reported in Section 3 correspond to the held-out test partition.

#### 2.3.2. Boundary Attention Mapper (BAM)

BAM [18] produces segmentation masks by fusing two sources of information from the trained EfficientNet-B7. The first source comprises 64 feature maps from the initial convolutional layer (each 300 × 300), which retain high spatial resolution and encode boundary and texture details. The second is the GradCAM heatmap computed from the final convolutional block (19 × 19 × 2560), which captures class-discriminative attention but at coarse resolution.

The algorithm selects a subset of the 64 first-layer channels whose average activation maximizes Pearson correlation with the upsampled GradCAM map. For a given image, this might be channels 38, 49, 54, and 57. Averaging the selected channels produces a single 300 × 300 visualization that combines the spatial precision of early features with the semantic selectivity of GradCAM. Adaptive thresholding converts this visualization into a binary segmentation mask (Figure 4, bottom). The practical advantage over raw GradCAM segmentation is resolution: BAM masks preserve fine boundary detail at the full input resolution rather than being constrained to the 19 × 19 grid of the last layer.

### 2.4. ABCD Feature Extraction

Once a lesion is segmented, its ABCD features are computed. A standardized alignment step precedes feature extraction, as illustrated in Figure 5.

**Figure 5.**
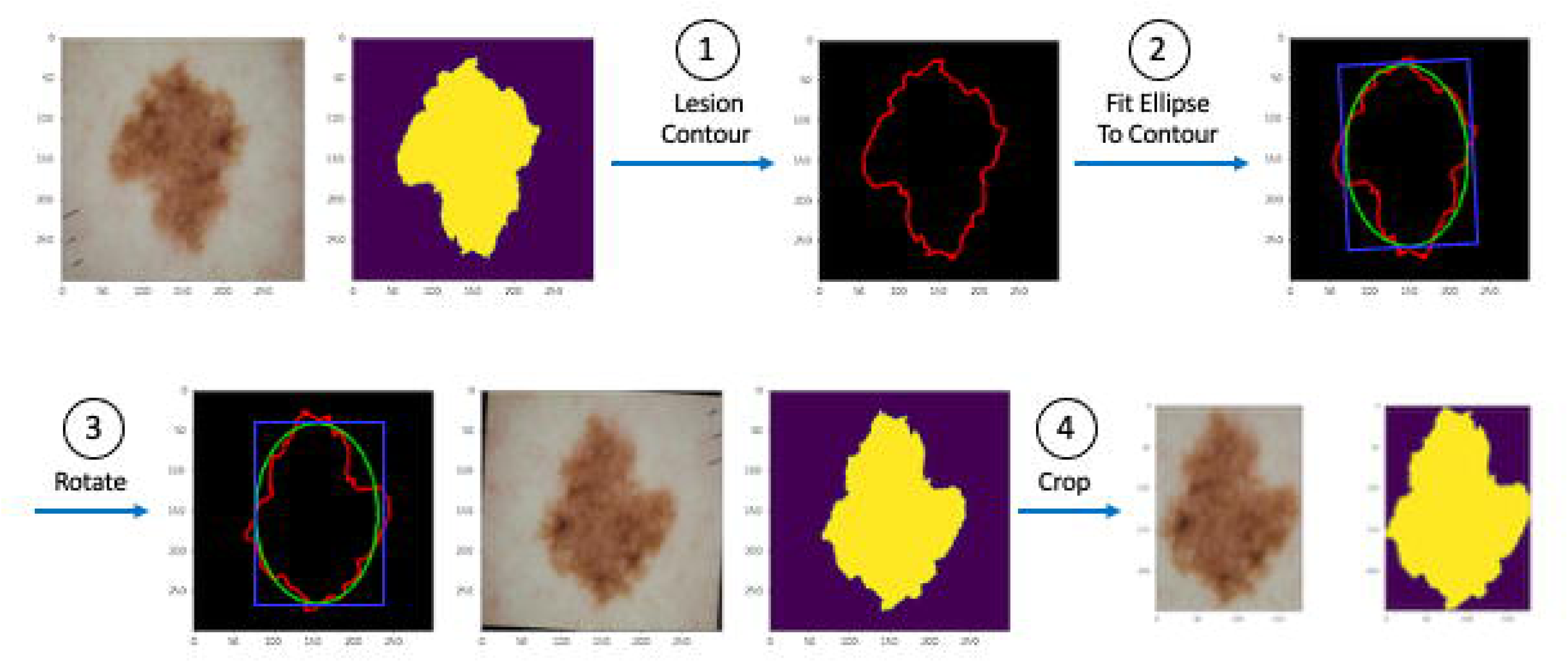
Pre-processing steps performed before measuring A, B, C, and D features for a lesion.

#### 2.4.1. Lesion Alignment and Cropping

The lesion’s external contour is extracted from the binary mask using *findContours* (external retrieval mode). An ellipse is fitted to the largest contour via *fitEllipse*, returning center coordinates, axes lengths, and rotation angle θ. The lesion image and mask are both rotated by θ to align the ellipse with the image axes, then re-binarized at a threshold of 128 to correct interpolation artifacts. The result is cropped to the lesion’s tight bounding box (Figure 5).

#### 2.4.2. Asymmetry (A)

Let *A*_T be the total number of lesion pixels in the cropped mask. The mask is split at its midpoint, first horizontally, then vertically. For each split, the lesion pixel counts in the two halves (*A*₁, *A*₂) yield an absolute difference:

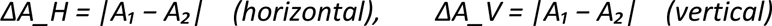

Asymmetry scores for each axis are:

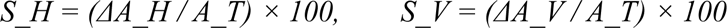

The final Asymmetry Index is max (S_H, S_V), ranging from 0 (perfect symmetry) to 100 (extreme asymmetry). Taking the maximum ensures the worst-case axis dominates. In the example shown in Figure 6, horizontal and vertical scores of 11.33 and 3.17 yield a final index of 11.33.

**Figure 6.**
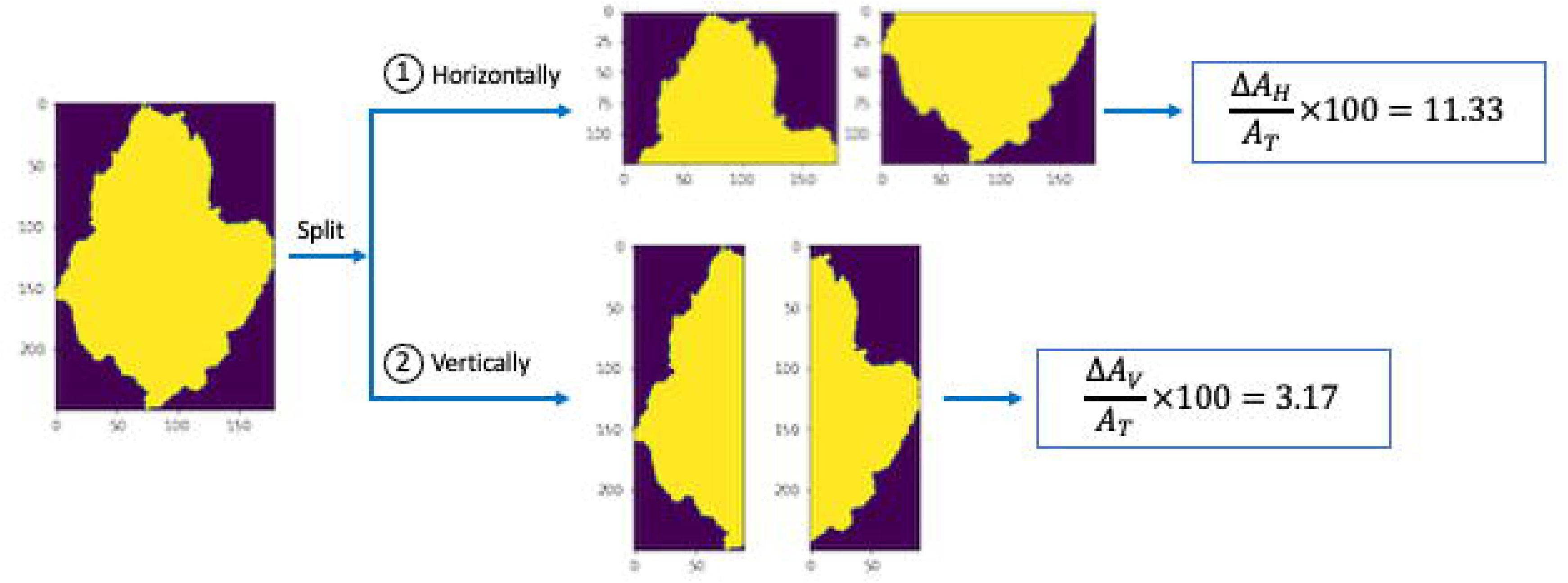
Measuring asymmetry for a lesion.

#### 2.4.3. Border Irregularity (B)

Border irregularity is measured by the Compactness Index, the isoperimetric quotient of the lesion contour:

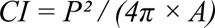

where *P* is the contour perimeter and *A* is the enclosed area. A perfect circle gives CI = 1. Jagged, notched, or angulated borders drive CI above 1 with no upper bound; higher values mean greater irregularity. Figure 7 shows a lesion with CI = 2.21 adjacent to a reference circle with CI = 1.

**Figure 7.**
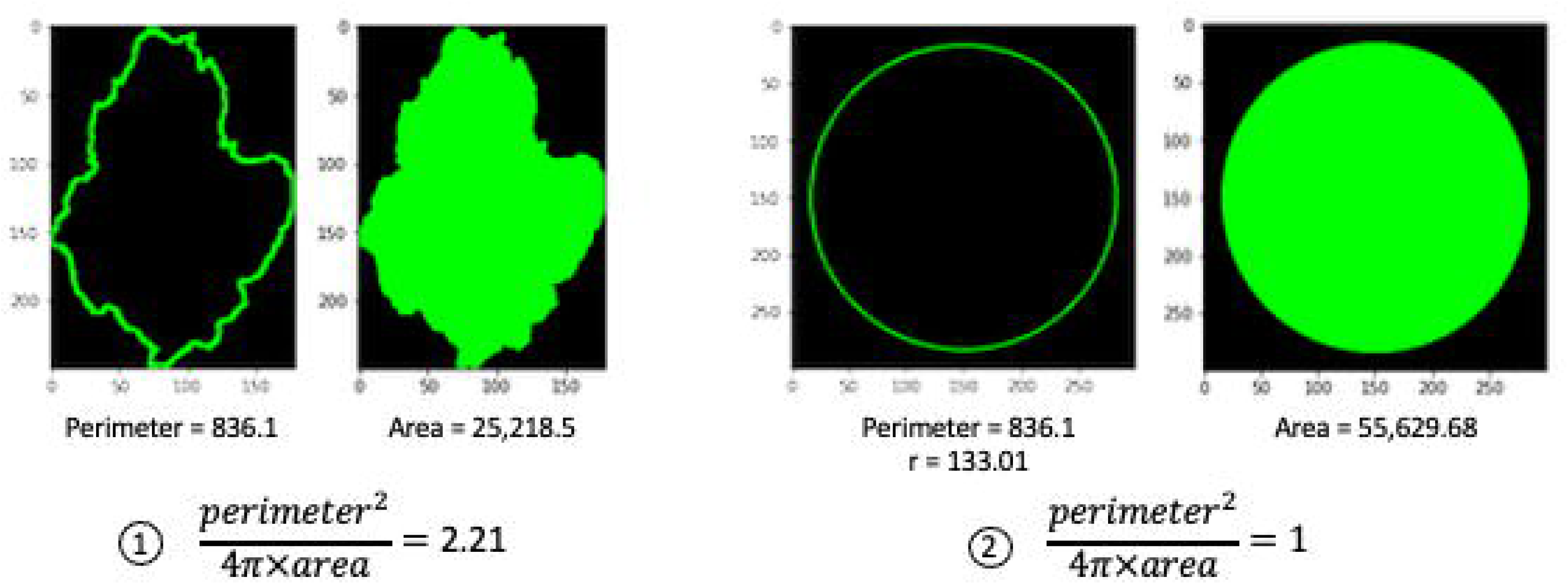
Measuring border irregularity for a lesion.

#### 2.4.4. Color Variegation (C)

Distinct colors within a lesion are counted by K-means clustering of the RGB pixel values, after excluding non-lesion pixels. Rather than the elbow method, we use a threshold-based stopping rule: K-means runs for *k* = 1 through 7, and clustering terminates when the sum of squared distances (SSD), scaled by 10⁻⁸, falls below 0.23.

That threshold was calibrated empirically on the PH2 dataset [19], which has expert-annotated color counts for 200 dermoscopic images. A sweep from 0.05 to 0.35 found that 0.23 minimizes the mean squared error between predicted and ground truth color counts (MSE = 1.425; Figure 9). Detected colors range from 1 to 10 per lesion, with higher values indicating greater variegation and correspondingly greater suspicion. Figure 8 illustrates the clustering process and resulting color pie charts for k = 1 through 4.

**Figure 8.**
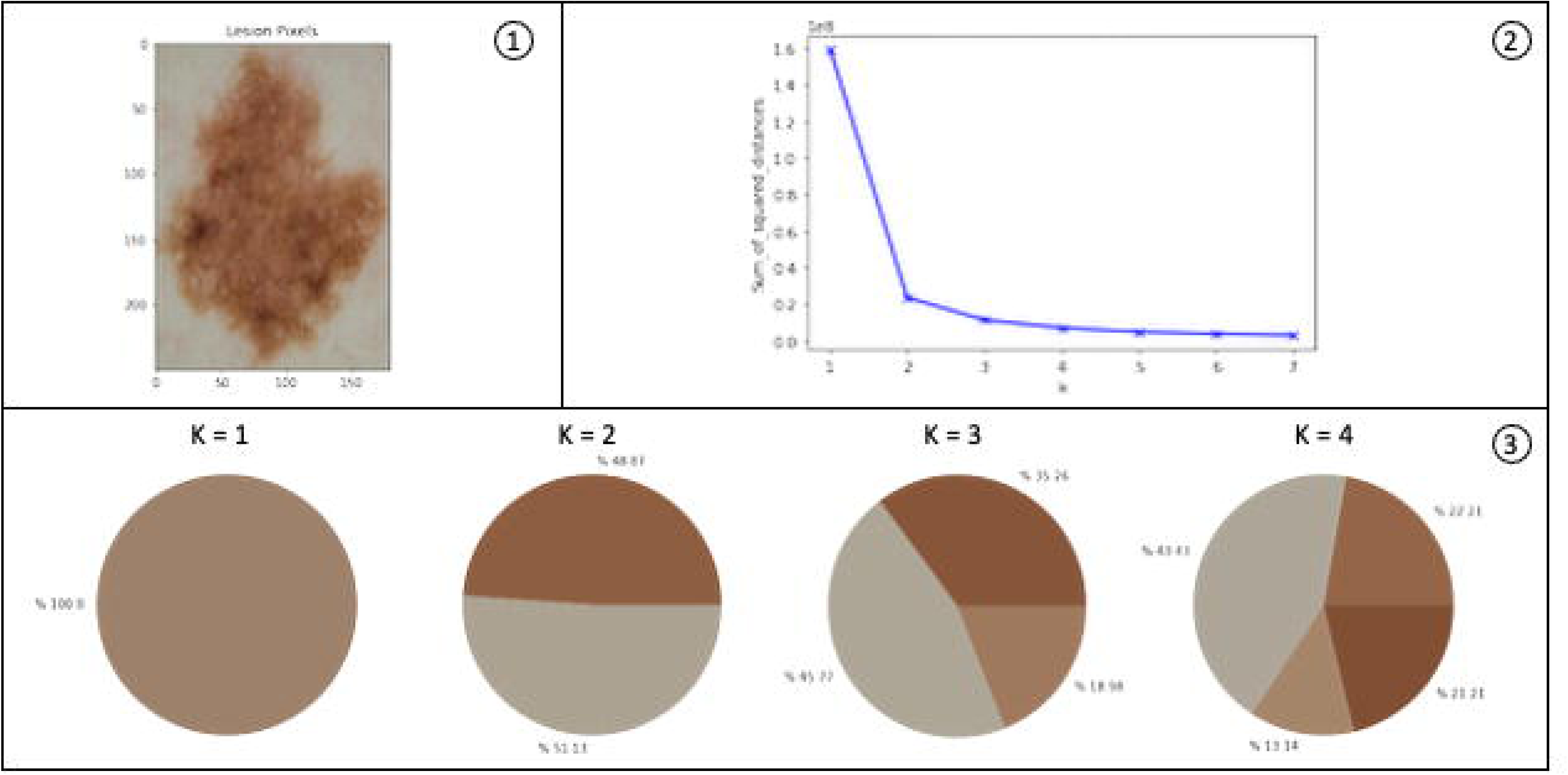
K-means clustering of lesion colors.

**Figure 9.**
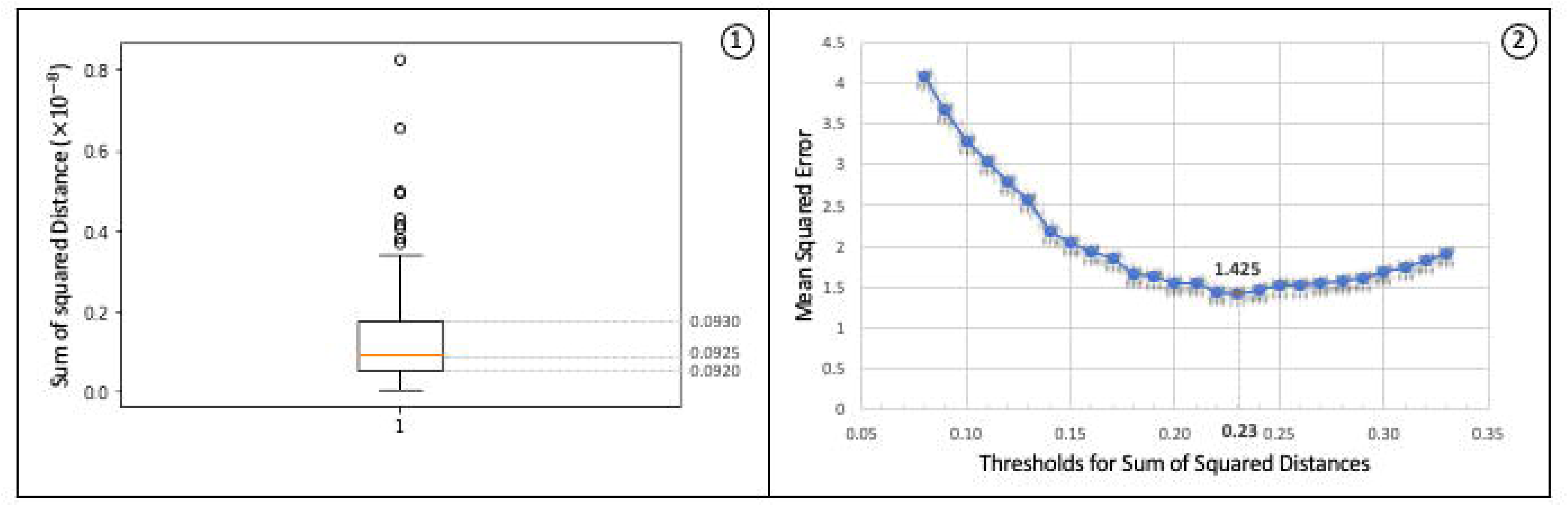
Color threshold optimization using the PH2 dataset.

#### 2.4.5. Diameter (D)

Diameter is the maximum Euclidean distance between any two points on the lesion contour:

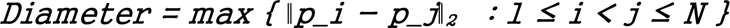

computed over all *N*(*N*−1)/2 pairs. The measurement is in pixels by default. If a fiducial marker of known dimensions is present, a pixel-to-millimeter ratio enables conversion to physical units. Clinically, diameters exceeding 6 mm raise concern (Figure 10).

**Figure 10.**
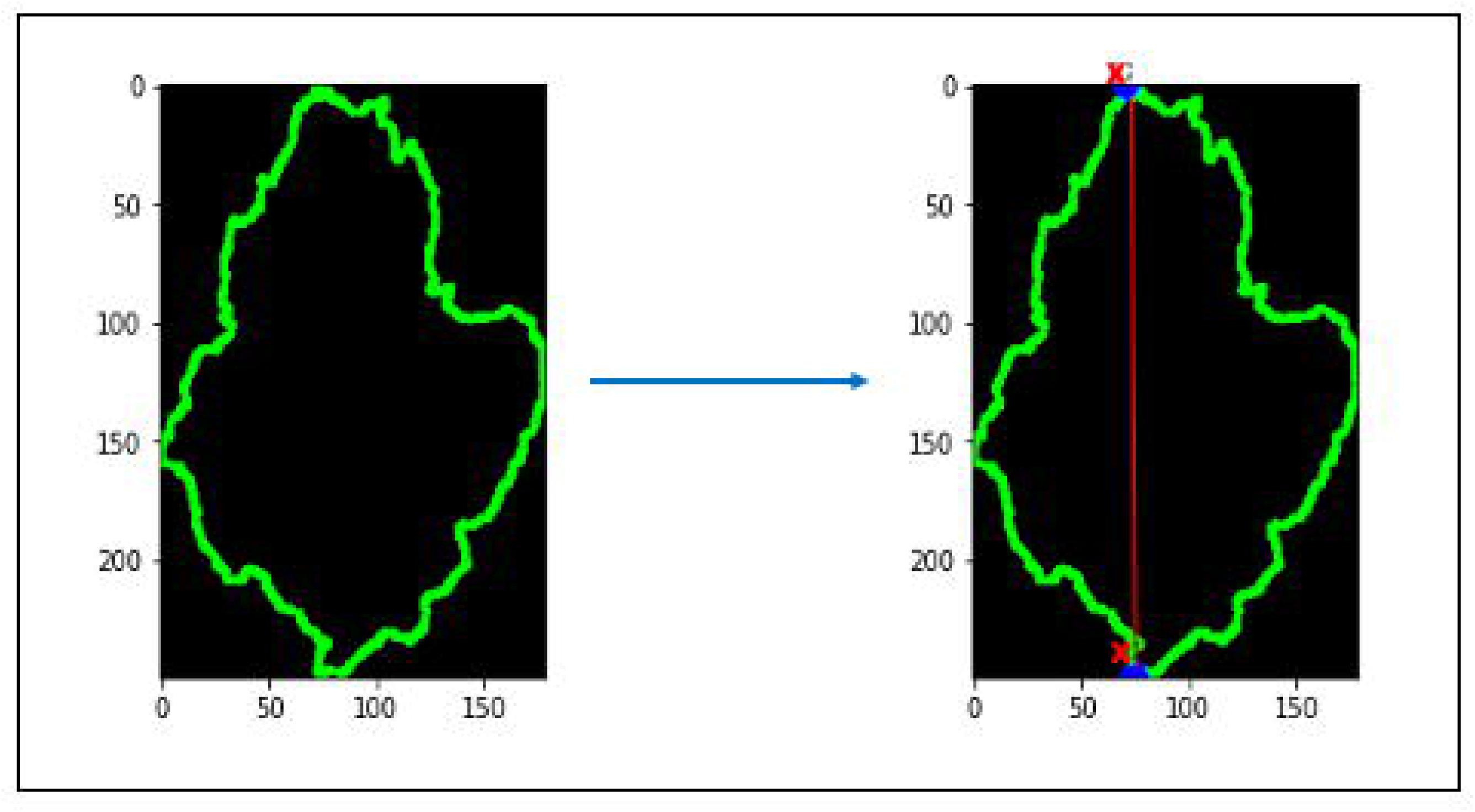
Measuring diameter for a lesion.

### 2.5. Output and Reporting

The pipeline produces a CSV file with one row per validated mole: mole identifier, horizontal and vertical asymmetry indices, final asymmetry index, compactness index, color count, and diameter in pixels. This output feeds directly into clinical triage workflows, longitudinal tracking, or downstream classifiers.

## 3. Experiments and Results

### 3.1. Lesion Segmentation Performance

BAM and conventional GradCAM segmentation were compared on the ISIC2017 dataset (2,750 dermoscopic images with expert-annotated masks). Four metrics were computed (pixel-level accuracy, sensitivity, specificity, and F-score) separately for melanoma, seborrheic keratosis, and other lesions overall.

BAM achieved 90.45% overall accuracy versus 87.78% for GradCAM (Table 1). The largest gain was in sensitivity: 86.06% versus 77.64%, an 8.42 percentage point difference. BAM captures more of the true lesion area while simultaneously being more selective about non-lesion pixels (specificity: 94.35% vs. 91.51%). The overall F-score improved from 75.02% to 79.55%.

**Table 1.** Segmentation Performance Comparison: BAM vs. GradCAM on ISIC2017 Dataset.

| Method / Class | Accuracy (%) | Sensitivity (%) | Specificity (%) | F-Score (%) |
| --- | --- | --- | --- | --- |
| GradCAM |  |  |  |  |
| Melanoma | 83.39 | 75.15 | 86.92 | 76.50 |
| Seborrheic Keratosis | 88.94 | 78.89 | 93.33 | 74.82 |
| Other | 91.02 | 78.88 | 94.27 | 73.73 |
| Overall | 87.78 | 77.64 | 91.51 | 75.02 |
| BAM (Proposed) |  |  |  |  |
| Melanoma | 87.81 | 81.97 | 94.15 | 82.42 |
| Seborrheic Keratosis | 90.04 | 83.46 | 94.56 | 77.12 |
| Other | 93.49 | 92.75 | 94.34 | 79.10 |
| Overall | 90.45 | 86.06 | 94.35 | 79.55 |
| Improvement (BAM over GradCAM) |  |  |  |  |
| Overall | +2.67 | +8.42 | +2.84 | +4.53 |

The melanoma class showed the most consequential improvement: accuracy increased from 83.39% to 87.81%, sensitivity from 75.15% to 81.97%, and F-score from 76.50% to 82.42%. These per-class gains matter because melanoma is the lesion type where segmentation errors are least tolerable; border irregularity and diameter are computed directly from the mask boundary.

Figure 11 shows the difference qualitatively. GradCAM heatmaps are diffuse and low-resolution, frequently bleeding beyond the true lesion edge or failing to cover the full lesion. BAM masks are sharp and closely match the ground truth, especially at borders.

**Figure 11.**
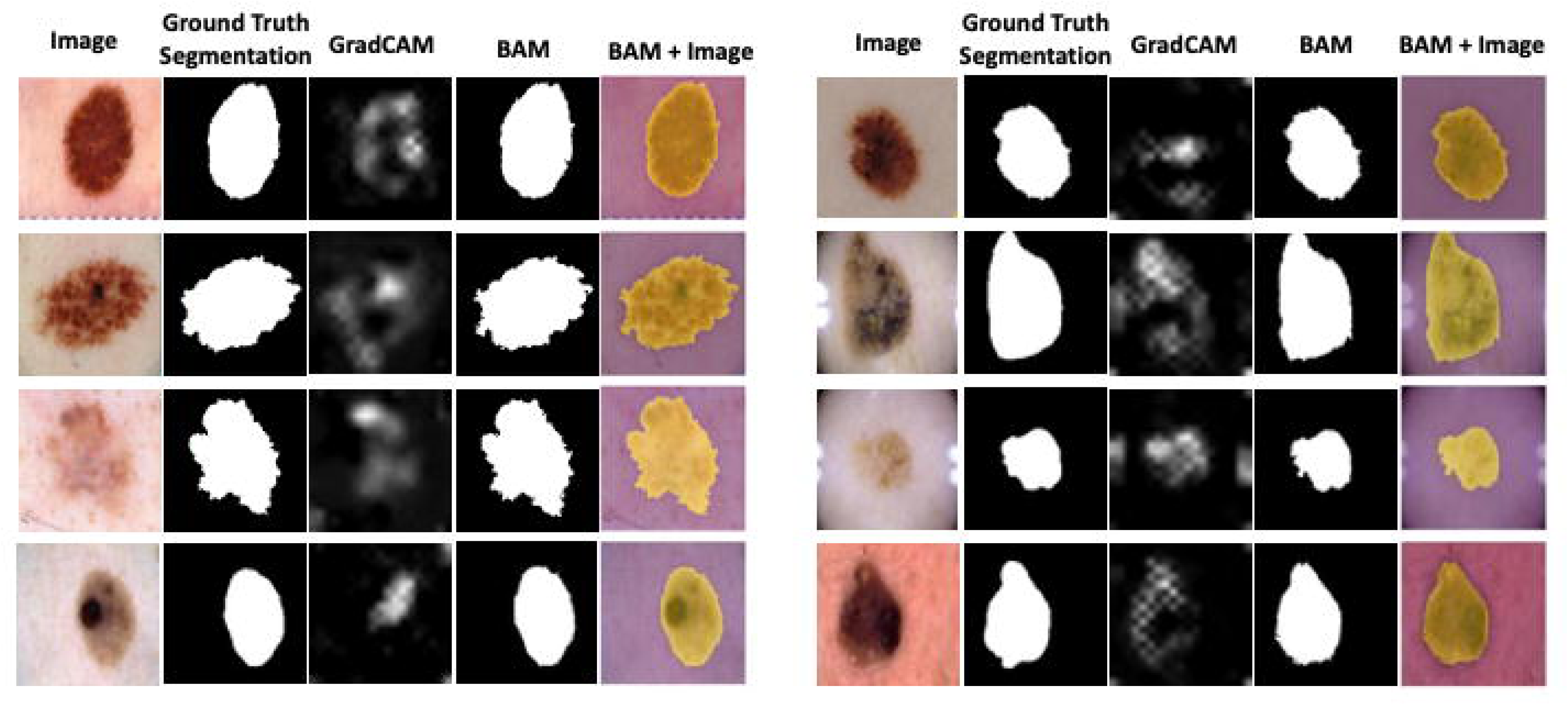
Visual comparison of segmentation methods on the IS1C2017 dataset.

We also compared BAM with two dedicated segmentation architectures, DeepLabv3 and Segment Anything Model (SAM) v2, on a smaller test subset. Both models were fine-tuned on the ISIC2017 training split. DeepLabv3, which uses atrous convolution and Atrous Spatial Pyramid Pooling (ASPP) to capture multi-scale context, and SAM v2, a transformer-based promptable segmentation model capable of zero-shot generalization, each produced Dice and pixel-level accuracy scores more than 5 percentage points below BAM on this test set. The gap is consistent with the design advantage BAM exploits: rather than learning segmentation from scratch, BAM extracts spatial information from a classifier already trained to attend to lesion-relevant features, effectively leveraging 36,000 training images’ worth of discriminative signal for a task neither DeepLabv3 nor SAM was optimized for in this domain without comparable data.

### 3.2. Lesion Classification Performance

The EfficientNet-B7 classifier was evaluated across all eight lesion categories. The confusion matrix and ROC curves are in Figure 12.

**Figure 12.**
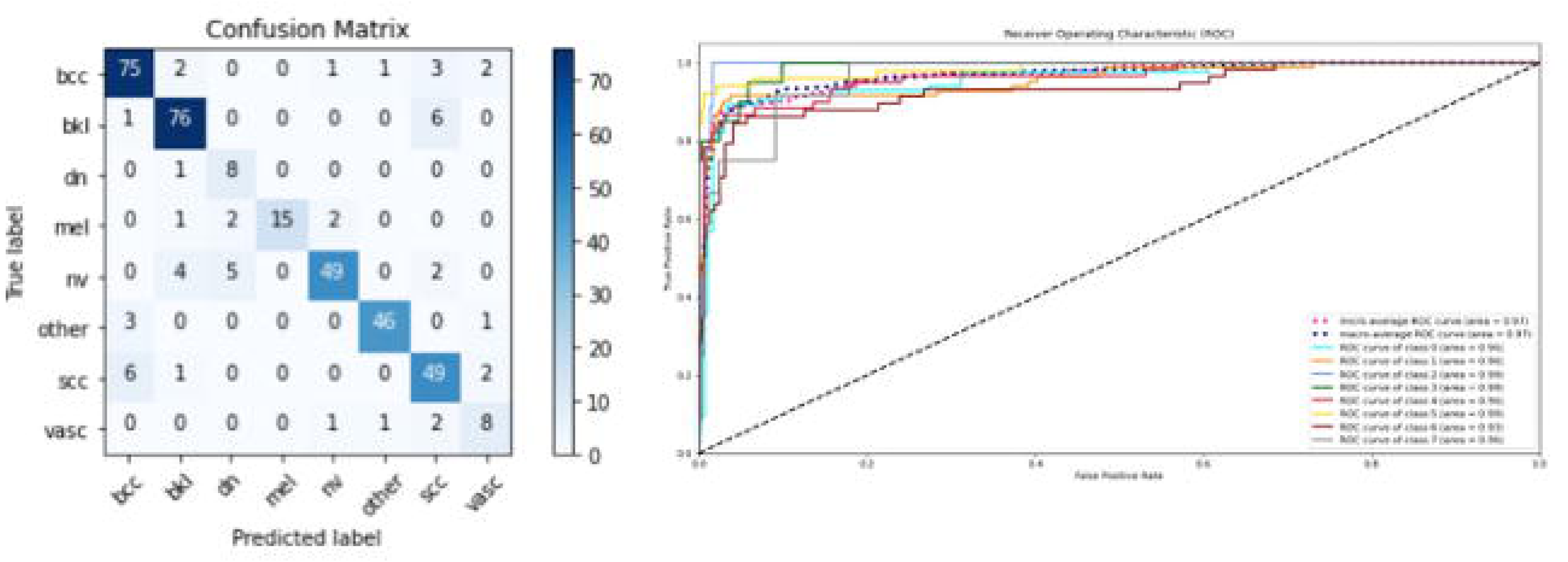
Classification performance of the EfficientNet-B7 lesion classifier.

Micro-average and macro-average AUC were both 0.97. Per-class AUC values: bcc 0.96, bkl 0.96, dn 0.99, mel 0.99, nv 0.96, other 0.99, scc 0.93, vasc 0.96. The confusion matrix shows clean separation for benign keratosis-like lesions (76/83), other lesions (46/50), and melanocytic nevi (49/60). The main points of confusion were SCC misclassified as BCC (6/58) and melanoma confused with dermatofibroma (2/20). The melanoma AUC of 0.99 indicates strong discrimination at the classification threshold that clinically matters most.

### 3.3. Color Feature Validation on PH2 Dataset

Color counting was validated on the PH2 dataset [19]: 200 dermoscopic images (80 common nevi, 80 atypical nevi, 40 melanomas) at 768 × 560 pixels, acquired with the Tuebinger Mole Analyzer from Fitzpatrick type II–III skin.

K-means clustering at the ground truth *k* for each PH2 image yielded a median SSD (scaled by 10⁻⁸) of 0.093, with an interquartile range of 0.092–0.093 and outliers to 0.8 (Figure 9, left). Sweeping the SSD threshold from 0.05 to 0.35 produced a U-shaped MSE curve; the minimum of 1.425 occurred at a threshold of 0.23 (Figure 9, right), which we adopted for all subsequent experiments.

### 3.4. Mole Detection and False Positive Removal

The detection pipeline was run on 20 wide-angle images (iPhone 14 Pro, 1080p) of backs and lower arms. Figure 3 shows a representative case: initial blob detection identifies a dense set of candidates, the brightness filter strips out false positives, and EDSR enhancement recovers boundary detail in the surviving crops. Processing was parallelized across images using *joblib*.

To obtain quantitative detection performance, we extended the evaluation to an additional 67 wide-angle images, each manually annotated with ground truth mole locations. Across this expanded set, the full detection pipeline (blob detection followed by false positive removal) achieved an F1 score of 86%. This indicates that the combination of adaptive thresholding, area-constrained blob detection, and brightness-based filtering provides a reasonable baseline for mole localization in consumer-grade wide-angle photographs, though there is clear room for improvement with learned detection models.

## 4. Discussion

### 4.1. Interpretation of Results

The central finding of this work is that BAM produces substantially better lesion segmentations than standard GradCAM on the ISIC2017 benchmark, and that this improvement is large enough to matter for downstream ABCD feature computation. The sensitivity gain was 8.42 percentage points overall, 6.82 for melanoma specifically. In clinical terms, it means the system delineates lesion boundaries more completely, which directly affects the accuracy of border irregularity and diameter measurements. A segmentation method that routinely under-segments by 20–25% of the true lesion area (as GradCAM does, based on its 75.15% melanoma sensitivity) will systematically underestimate lesion perimeter and undercount border notching. BAM’s improvement closes much of that gap.

The mechanism behind this improvement is straightforward. GradCAM operates at the resolution of the final convolutional layer (19 × 19 pixels in EfficientNet-B7) and must be upsampled to the input resolution, which inevitably blurs boundary detail. BAM sidesteps this limitation by using GradCAM only as a semantic guide: it identifies which first-layer channels (at 300 × 300) carry lesion-relevant information and averages them to produce a high-resolution map. The result preserves the spatial precision of early network layers while retaining the class-discriminative signal from the final layer. This is not a new insight, since the tension between resolution and semantics in CNN feature hierarchies is well known, but BAM provides a practical and effective resolution.

The classification results reinforce confidence in the backbone network. An AUC of 0.99 for melanoma means the EfficientNet-B7 classifier can reliably separate melanoma from non-melanoma at clinically useful operating points. This is relevant because BAM depends on the classifier’s learned representations; if the classifier were unreliable, the saliency maps it generates would be unreliable too. The strong classification performance suggests that the features BAM draws on are genuinely disease-relevant, not artifacts of overfitting.

### 4.2. Clinical Relevance and Workflow Integration

The practical value of this pipeline lies in what it automates and what it does not. It automates the tedious, time-consuming parts of mole mapping (detecting individual lesions in wide-angle photos, segmenting them, and computing standardized ABCD features) while leaving diagnostic interpretation to the clinician. The output is a structured table of feature scores, not a diagnosis. A dermatologist can scan the table, sort by asymmetry or compactness index, and quickly identify the lesions that warrant closer inspection or biopsy. This preserves clinical judgment while reducing the bottleneck of manual lesion-by-lesion documentation.

The system’s compatibility with smartphone cameras is clinically relevant. Dermoscopy equipment produces higher-quality images, but is expensive, requires training, and is unavailable in most primary care settings and essentially all home-monitoring contexts. The 14.1-minute baseline exam time reported by Nazari and Garcia [9] describes a process that a primary care physician in a 15-minute appointment slot simply cannot perform.

### 4.3. Comparison with Existing Systems

Most existing CAD systems for melanoma operate on single dermoscopic images: one lesion per image, pre-cropped, under controlled lighting. Our pipeline addresses a different and arguably harder problem: starting from a wide-angle photograph of an entire body region and working down to per-lesion feature scores. This end-to-end scope introduces challenges (mole detection, false positive filtering, resolution recovery) that dermoscopic systems do not face, but it also provides a more complete clinical workflow.

The segmentation accuracy of BAM (90.45% overall on ISIC2017) is competitive with dedicated segmentation networks trained specifically for the task. Our direct comparison with DeepLabv3 and SAM v2, both fine-tuned on the same data, showed BAM outperforming each by more than 5 percentage points in Dice and pixel-level accuracy. This is striking because BAM is not a segmentation model; it is a post-hoc extraction of spatial information from a classifier. The practical advantage for deployment is that a single EfficientNet-B7 serves double duty as both classifier and segmentation backbone, reducing model complexity and inference overhead compared to pipelines that require separate networks for each task.

The color quantification approach also differs from prior work, which typically uses the elbow method or silhouette scores to determine *k* in K-means clustering. These generic methods are not calibrated to dermatological color categories. Our threshold-based stopping rule, calibrated against PH2 ground truth, is domain-specific and produces color counts with a mean squared error of 1.425, not perfect, but grounded in clinical color annotations rather than statistical heuristics.

### 4.4. Limitations

Several limitations should be stated plainly. First, while the mole detection pipeline achieved an F1 score of 86% across 87 wide-angle images (20 initial plus 67 additional), the evaluation dataset remains modest in size and was captured predominantly with a single device (iPhone 14 Pro). Detection performance on images from other smartphones, under different lighting conditions, or on skin types not well represented in the test set may differ. An 86% F1 is workable for a screening tool, since most missed moles will be caught at the next visit, but a system intended for real-world clinical deployment would need validation on a larger, multi-device, multi-institution dataset.

Second, the PH2 dataset used to calibrate the color threshold contains only 200 images, all from Fitzpatrick skin types II and III. Whether the threshold of 0.23 generalizes to darker skin tones is an open question. K-means clustering on skin lesion pixels is sensitive to the baseline skin color, and recalibration on a dataset spanning Fitzpatrick types I through VI is needed.

Third, the current pipeline treats each image independently. It does not track lesions across visits or flag changes over time, the “Evolution” component of the ABCDE framework. Longitudinal comparison is arguably the most powerful clinical signal for melanoma detection, and its absence is a meaningful clinical gap.

Fourth, the automated ABCD feature scores have not yet been validated against expert clinical ratings or correlated with histopathological diagnosis. The features are computed according to established dermatoscopic definitions, but whether the pipeline’s asymmetry index of 11.33 means the same thing to the algorithm as it does to a dermatologist looking at the same lesion is an empirical question. A clinical correlation study is currently underway, with two board-certified dermatologists and two surgeons independently scoring a subset of lesions to establish inter-rater agreement and algorithm-to-clinician concordance. The results of that study will be reported separately.

Fifth, the diameter measurement is reported in pixels. Without a fiducial marker or known camera-to-skin distance, there is no way to convert to millimeters. The clinical 6 mm threshold for diameter cannot be applied in absolute terms without physical calibration, which the current system does not provide automatically.

Finally, the false positive filter uses a fixed amplification factor (*r* = 1.2 or 1.3) derived from heuristic brightness thresholds. This works adequately on the images tested, but the rule is brittle; it could fail on images with unusual lighting, very dark skin, or lesions that are lighter than the surrounding skin (e.g., amelanotic melanoma). A learned false positive classifier, trained on labeled examples, would likely be more reliable.

### 4.5. Future Directions

Three extensions would most improve the system. First, completing the ongoing clinical correlation study: the results from the two dermatologists and two surgeons currently evaluating the pipeline’s ABCD scores will establish whether automated feature values meaningfully correspond to expert clinical assessments. That study is the most immediate next step. Second, adding temporal tracking: aligning lesion positions across successive photographs of the same patient to detect evolving moles. This is technically feasible through landmark-based image registration and would bring the system closer to a full ABCDE implementation. Third, validating the pipeline on a multi-center dataset spanning diverse skin types, camera hardware, and lighting conditions, particularly recalibrating the color threshold across Fitzpatrick types I through VI. The heuristic components of the pipeline (the brightness-based false positive filter, the threshold-based color stopping rule) are its weakest links and the most natural targets for replacement with learned models trained on annotated data.

## Data Availability

All data produced in the present work are contained in the manuscript

